# Prevalence of, and knowledge about intestinal helminths among pregnant women at Adidome and Battor district Hospitals

**DOI:** 10.1101/2023.03.23.23287608

**Authors:** Charity Ahiabor, Naa Adjeley Frempong, Atikatou Mama, Kwadwo A. Kusi, William Anyan, Michael F. Ofori, Bright Adu, Yvonne Ashong, Bernard W. Lawson, Abraham K. Anang, Nicaise T. Ndam, David Courtin

## Abstract

**Background:** Soil Transmitted helminths and schistosomiasis may have implications for pregnancy outcomes especially, in rural communities. In Adidome (a rural community) and Battor (a semi-rural community), soil and water contact activities expose inhabitants to helminth infections. There is, however, limited information on the prevalence and determinants among pregnant women in these areas. The present study was conducted to access the prevalence, knowledge and perceptions about helminthiasis among pregnant women accessing obstetric care at Adidome Government Hospital and Battor Catholic Hospital in the Volta region of Ghana.

**Methodology/Principal Findings:** A cross-sectional survey was conducted by recruiting 1,295 pregnant women out of which only 616 representing 47.5% provided stool samples for analysis. Sociodemographic characteristics and participant knowledge on helminth infection transmission, symptoms and prevention were collected by questionnaire and analyzed using STATA. Preserved stool specimen were processed and examined for helminth eggs by Kato Katz, and genomic DNA extracted from aliquots, was tested for *S. mansoni* and *N. americanus*. Prevalence of helminths and participant knowledge were expressed in proportions. Chi-square and Fisher’s exact test were used to show association at P < 0.05 significant level. Intestinal helminth infections found among participants at ANC were *T. trichiura* (0.4%), *N. americanus* (0.4%) and *S. mansoni* (0.4%). At delivery, a PCR prevalence of 5% was observed for *S. mansoni*. A high proportion of study participants, 82.5% in Adidome and 87.1% in Battor do not take dewormers on regular basis. Also, a high proportion of participants did not receive any dewormer prior to sample collection. Although knowledge on helminth transmission, risks and prevention were low, levels of prevalence of infection with helminths were also low.

**Conclusion/Significance:** Intensifying health education as community-based interventions is necessary for the total and effective control and elimination of schistosomiasis and STH in the study area.

**Author summary:** Adidome and Battor are two communities sited close to the Volta Lake which is known to be infested with intermediate hosts for schistosomiasis. Consequently, there has been continued efforts towards control by mass deworming especially among pre-school and school-going children. Despite these control efforts, infections persist as older age groups including pregnant women are often left out of such programs. The fertile soil in the Volta basin encourages farming activities which further expose the people to soil transmitted helminths. There is therefore a need to evaluate community knowledge on helminth infection and precautions to be taken to improve on the control efforts in the areas. The study selected two district Hospitals (a private Hospital in Battor and a government Hospital in Adidome) as study sites. Stool samples collected from pregnant women reporting for the first antenatal care and for delivery were examined for the presence of helminths. Knowledge about helminths among participants were collated by questionnaire. Although prevalences of helminth infections were low, knowledge about helminth infections were also low. However, participants from Battor appear more knowledgeable about helminth infections than those in Adidome. Our study therefore suggests intensive public health education as an intervention for control.

## Introduction

Schistosomiasis and Soil Transmitted Helminthiasis (STH) are parasitic diseases of medical and public health importance in many parts of the world. Classified among the neglected tropical diseases, they are prevalent in populations characterized **by** poverty and poor sanitation. STHs infect more than 1.5 billion people worldwide (1) accounting for nearly 3.3 million disability-adjusted life years [DALYs]. Despite the goals set at the 54th World Health Assembly resolution (2), the number of people affected by these neglected diseases remains very high (3, 4).

In Ghana, the predominant STHs are *Necator americanus, Ascaris lumbricoides and Trichuris trichiura* (5). Transmission is either oral or by direct skin contact with the infective stages. Significant morbidities have been attributed to STH infections, including anemia, malnutrition, growth retardation and low intellectual ability (6). For pregnant women, these complications might have implications for pregnancy outcomes and neonatal health (3).

Two forms of schistosomiasis are observed in Ghana, ie urinary schistosomiasis caused by *Schistosoma haematobium* and intestinal schistosomiasis caused by *S. mansoni*. Transmission is by contact with contaminated freshwater body which harbours the specific snail intermediate hosts. People become infected during routine activities which bring them into contact with infected water bodies. In endemic communities, concurrent infections of the two forms of the disease have been reported (7). Symptoms include fever, joint and abdominal pains, bloody diarrhea and hematuria and the infections are also known to contribute to the burden of anaemia in pregnancy (8)

Generally, higher prevalence of helminth infections are observed in younger people living in communities with inadequate sanitation, overcrowding, and low socioeconomic status (4,5) than in their older counterparts. Chronic STH infections tend to be asymptomatic (5) especially in low intensities and can prolong for a lifetime into adulthood.

In recent years, there has been a growing interest in the effect of helminth infections on pregnancy and pregnancy outcomes. Chronic helminth infection is reported to significantly increase maternal risks for future parasitic infections and adverse pregnancy outcomes. Infants born to malnourished mothers are often delivered prematurely, with low birth weight, and may have poor growth and development throughout childhood. (9,10).

Even though there is global commitment to implementing helminth control programs, the primary target population is usually school children. Older age groups including pregnant women are often left out despite the risk of serving as reservoirs for reinfection within the communities (11). Deworming with single dose 400mg albendazole or 500mg mebendazole has been used as public health intervention for pregnant women after the first trimester especially in communities where baseline prevalences of ≤20% have been observed for Hookworm and *T. trichiura* (12).

Battor-Dugame and Mafi-Adidome are two district capitals of the North Tongu and Central Tongu districts, respectively, which share a common boundary, thereby encouraging trade among them. The two communities are situated close to the Volta Lake which was created after the damming of the Volta River and consequently, has become infested with the snail intermediate hosts for schistosomiasis. The lake serves as a source of potable water for many households. Farming activities in the Volta basin along the Lake expose the people to STHs while water contact activities also expose them to schistosomiasis. Information on helminth infections among pregnant women in the study area is limited, following the continued focus on school children for helminth control (13,14).

Adidome Government Hospital (AGH) and Battor Catholic Hospital (BCH) are the main referral centers in Mafi-Adidome and Battor-Dugame, respectively. Stool examinations in these hospitals are performed only upon the request of the medical officer in charge and not on routine basis. Consequently, the needed treatment for asymptomatic pregnant patients may be overlooked. The WHO target for the elimination of schistosomiasis and soil-transmitted helminths by 2030 warrants a clear understanding of community knowledge about these infections.

The **aim** of this study, therefore, was to determine prevalence of STH and *S. mansoni* and risk factors in relation to local knowledge on symptoms and prevention amongst pregnant women reporting at Adidome Government Hospital and Battor Catholic Hospital for obstetric care.

The specific objectives were to:

1. Determine prevalence of STH and *S. mansoni*.
2. Evaluate the deworming history among study participants.
3. Estimate local knowledge on causes, symptoms and prevention of helminth infections.
4. Show knowledge about helminth infections across sociodemographic characteristics.

## Methods

### Study area and study site

Battor-Dugame (North Tongu District capital) and Mafi-Adidome (Central Tongu District capital) have tropical climate which is favorable for farming, a major occupation for 73.5% of households in North Tongu and 78.4% in Central Tongu. There are two rainy seasons: a major season from mid-April to early July and a minor season from September to November. Average annual rainfall varies between 900 mm to 1100 mm with 50% occurring in the major season. Daily minimum and maximum temperatures range between 22oC and 33oC with average humidity around 80% (15,16). The natural vegetation is savannah grassland. Aside crop farming, some households also engage in fish farming, livestock rearing and petty trading. Transportation in the north and central Tongu districts is by motorcycles, taxies, small buses and private cars. People living on the banks of the lake also use small canoes for navigation. Accommodation in Battor-Dugame and in Mafi-Adidome ranges from large extended family settlements to smaller nuclear family houses. Toilet facilities range from privately owned to public pit latrines, KVIPs and water closets. Some residents however, resort to open defecation in the surrounding bushes due to the absence of these facilities within their households.

Battor-Dugame has a semi-urban structure with a literacy rate of 74% (ie 51.7% male and 48.4% female) (15) while Mafi-Adidome has sparse or scattered settlements with small clusters of households near the central market. The literacy rate is 72% ie 52% male and 48% female (16). The catchment area for AGH includes neighboring towns and small communities such as Mepe, Volo, Mafi-Kumase, Bakpa, Battor etc, while BCH also receives patients from adjoining towns and communities such as Mepe, Osudoku, Ada East and West, Volo, Abutia, Mafi-Adidome etc.

### Study design and participant recruitment

This is a cross sectional survey targeting pregnant women reporting at two health facilities for their first ANC visit and for delivery. Stool samples were collected from consenting pregnant women during the first ANC attendance and as they report for delivery, between November 2016 and March 2019. Participants who could not provide samples the same day were asked to bring their samples the following day. Information about sociodemographic characteristics and knowledge (or myths) on helminth infection transmission, prevention, and symptoms were collected by questionnaire interviews. Aliquots of the stool samples were preserved at 4oC for Kato-Katz detection and at -20oC for PCR.

### Sample size calculation

Sample size was calculated using a confidence interval of 95% and an estimated prevalence of 17% (17). The minimum sample size ‘n’ was calculated by the equation: 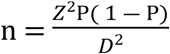 (18), where ‘D’ is the margin of error (5%), ‘n’ is the minimum sample size, ‘P’ is the estimated prevalence and ‘Z’ is the standard normal deviate that corresponds to the 95% confidence interval (16). Therefore, the minimum sample size required was 216 per study area.

### Ethical considerations

Ethical and scientific approvals for the study were obtained from the Ghana Health Service Ethical Review Committee (GHS_ERC06/06/16) and the Institutional Review Board of the Noguchi Memorial Institute for Medical Research (NMIMR-IRB CPN 071/15-16 amend. 2017). Participants who agreed to be part of the study and signed the consent form were assigned unique IDs. Parental consent was obtained on behalf of participating minors (<18 years) who also gave their assent.

### Stool analysis

Two Kato-Katz thick smears were prepared from each stool sample and observed microscopically for helminth ova. Genomic DNA was extracted from aliquots of the stool samples using the Qiagen stool DNA kit according to the manufacturer’s protocol after which species-specific primers were used to detect *Necator americanus* and *S. mansoni* by PCR. Optimization of PCR for *Ascaris* and *Trichuris* was not performed due to limited resource.

### Data capture and analysis

Knowledge on helminth infection acquisition, symptoms, prevention and associated risks for the mother and for the baby were obtained by questionnaire, analyzed and expressed in proportions. Chi-square test and Fisher’s exact test were used to show correlations between categorical variables and the outcomes at p<0.05 significance level.

## Results

### Sociodemographic characters

A total of 1,295 pregnant women were interviewed. However, only 616 pregnant women provided stool samples for analysis, comprising 249 participants from AGH and 367 participants from BCH. Mean ages of participants were 25.9(±6.5) years at AGH and 27.4(±6.0) years at BCH (t = -3.46, p<0.001). Most of the participants were basic school (primary and junior high school) leavers. The proportion of primary school leavers were 24.6% at AGH and 20.5% at BCH. While Junior High School (JHS) and Senior High School (SHS) leavers were 53.0% and 10.6% respectively at AGH and 43.4% and 14.0% respectively at BCH. Only a small proportion had completed tertiary level education. These were 1.6% at AGH and 10.8% at BCH (*X*^*2*^(4) =37.96, p<0.001).

### Prevalence of helminth infections

Species of helminths observed were *Schistosoma mansoni, Trichuris trichiura* and *Necator americanus*. Very low prevalences of 0.4% and 0.6% was recorded for *S. mansoni* by microscopy among ANC participants in Adidome and Battor respectively and this was confirmed by PCR at 0.4% and 1.4% respectively. STH infections were only observed among ANC participants. A microscopy prevalence of 0.4% for *T. trichiura* was recorded at Adidome ANC and a PCR prevalence of 0.4% for *N. americanus* was recorded at Battor ANC. Among participants at delivery, a PCR prevalence of 5% was recorded for *S. mansoni* in Battor.

### Deworming history, worm infection and knowledge about dewormers

Table 2 is the summary of the responses to questions on the deworming habits among ANC participants. Majority of them, comprising 82% in Adidome and 87% in Battor, do not regularly take dewormers. A Chi-square test however, showed a statistically significant difference (p<0.001) in regular deworming across the two health facilities. For those who regularly take dewormers, a proportion of 13.1% was observed in Adidome and 12.6% in Battor. Nearly all the participants (100% in Adidome and 99% in Battor) did not receive dewormers prior to stool sample collection. Also, whereas 29% of participants in Battor did not take dewormers because in their opinion, it was too early in the pregnancy, 89% in Adidome did not take dewormers because the medicines were not available in the health facility (*X*^*2*^(4) =634.48, p<0.001).

**Table 1:** Prevalence of STH and *S. mansoni*.

| Study Site | Microscopy (n, %) |  |  | PCR (n, %) |  |
| --- | --- | --- | --- | --- | --- |
|  | Dept/unit | *S. mansoni* | STH | *S. mansoni* | STH |
| AGH | ANC | 1(0.4%) | 1(0.4%) | 1(0.4%) | 0% |
|  | Delivery | 0% | 0% | 0% | 0% |
| BCH | ANC | 2(0.6%) | 0% | 4(1.4%) | 1(0.4%) |
|  | Delivery | 0% | 0% | 5(5.0%) | 0% |

AGH=Adidome Government Hospital, BCH= Battor Catholic Hospital, STH= Soil Transmitted Helminth

**Table 2:** Deworming habits and worm infection at ANC.

|  | ANC Health facility,<br>N (%) |  | Test statistic | P-value |
| --- | --- | --- | --- | --- |
|  | Adidome | Battor |  |  |
| Carry out regular deworming | | | $X^2(2)=14.75$ | $p < 0.001$ |
| No | 372 (82.5) | 331 (87.1) |  |  |
| Yes | 59 (13.1) | 48 (12.6) |  |  |
| Don't know | 20 (4.4) | 1 (0.3) |  |  |
| Had worms during current pregnancy | | | $X^2(2)=37.89$ | $p < 0.001$ |
| No | 435 (96.5) | 320 (84.2) |  |  |
| Yes | 1 (0.2) | 1 (0.3) |  |  |
| Don't know | 15 (3.3) | 59 (15.5) |  |  |
| Received dewormer today | | | $X^2(1)=4.77$ | $p = 0.043^f$ |
| No | 451 (100.0) | 376 (99.0) |  |  |
| Yes | 0 (0.0) | 4 (1.0) |  |  |
| Reason for no dewormer today | | | $X^2(4)=634.48$ | $p < 0.001$ |
| Too early in pregnancy | 22 (4.9) | 109 (29.0) |  |  |
| Not available in facility | 402 (89.1) | 6 (1.6) |  |  |
| Not offered | 19 (4.2) | 162 (43.1) |  |  |
| Not tested for worms | 6 (1.3) | 19 (5.1) |  |  |
| Other reasons | 2 (0.4) | 80 (21.3) |  |  |
| Total | 451 (100.0) | 380 (100.0) |  |  |

$X^2(df)$ = Chi-square (degrees of freedom), $f$ = Fisher's exact p-value.

Deworming habits and knowledge of dewormers among participants at delivery is presented in Table 3. Here too, a high percentage of participants reportedly did not take any dewormers during the pregnancy and they were 71.4% in Adidome and 87.1% in Battor. For those who took dewormers, 55.8% in Adidome and 61.9% in Battor took Albendazole, whiles 14% in Adidome and 19.1% in Battor took Mebendazole. Few participants however could not tell whether they had taken dewormers or not. Only a small proportion of participants (3% in Adidome and 1.2% in Battor) showed knowledge of herbal preparations against intestinal worm infections (p=0.343).

**Table 3:** Deworming history and knowledge about dewormers at delivery.

|  | Delivery Health facility,<br>N (%) |  | Test statistic | P-value |
| --- | --- | --- | --- | --- |
|  | Adidome | Battor |  |  |
| Taken dewormer during current pregnancy: | | | $\chi^2(1)=14.67$ | $p<0.001$ |
| No | 215 (71.4) | 142 (87.1) |  |  |
| Yes | 86 (28.6) | 21 (12.9) |  |  |
| Type of dewormer taken: | | | $\chi^2(3)=1.40$ | $p=0.706$ |
| Albendazole | 48 (55.8) | 13 (61.9) |  |  |
| Mebendazole | 12 (14.0) | 4 (19.1) |  |  |
| Don't know | 24 (27.9) | 4 (19.1) |  |  |
| Other | 2 (2.3) | 0 (0.0) |  |  |
| Know herbal preparation used to treat worms | | | $\chi^2(1)=1.42$ | $p=0.343^f$ |
| No | 292 (97.0) | 161 (98.8) |  |  |
| Yes | 9 (3.0) | 2 (1.2) |  |  |
| Can 0-6 month old babies get worm infestations? | | | $\chi^2(2)=4.90$ | $p=0.086$ |
| No | 46 (15.3) | 30 (18.4) |  |  |
| Yes | 85 (28.2) | 31 (19.0) |  |  |
| Don't know | 170 (56.5) | 102 (62.6) |  |  |
| Total | 301 (100.0) | 163 (100.0) |  |  |

$\chi^2(df)$ = Chi-square (degrees of freedom), $f$ = Fisher's exact p-value.

### Knowledge on helminthiasis

The level of participants’ knowledge about helminth infection estimated as the number of correct answers to questions and expressed as a percentage of the highest possible score is presented in Table 4. Knowledge levels were categorised into ‘Low’ (0-33.33%), ‘Medium’ (33.34-66.66%) and ‘High’ (66.67-100%). Very high percentages of participants (both at ANC and delivery) showed ‘Low knowledge’. Average score among ANC participants were 17.7% (±10.8) in Adidome and 26.0% (±15.8) in Battor (*X*^*2*^(2) =76.54, p<0.001). At delivery, average scores were 19.4% (±13.3) and 22.2% (±16.1) in Adidome and Battor respectively (*X*^*2*^(1) =1.19, p=0.276).

**Table 4:** Level of knowledge of helminthiasis.

|  | ANC Health facility,<br>N (%) |  | Test statistic<br>P-value | Delivery Health facility,<br>N (%) |  | Test statistic<br>P-value |
| --- | --- | --- | --- | --- | --- | --- |
|  | Adidome | Battor |  | Adidome | Battor |  |
| Knowledge level | | | $X^2(2)=76.54$<br>$p<0.001^f$ | | | $X^2(1)=1.19$<br>$p=0.276$ |
| Low | 430 (95.3) | 281 (74.0) |  | 242 (80.4) | 124 (76.1) |  |
| Medium | 21 (4.7) | 98 (25.8) |  | 59 (19.6) | 39 (23.9) |  |
| High | 0 (0.0) | 1 (0.3) |  | 0 (0.0) | 0 (0.0) |  |
| Total | 451(100.0) | 380(100.0) |  | 301(100.0) | 163(100.0) |  |
| Mean (SD) | 17.7 (10.8) | 26.0 (15.8) |  | 19.4 (13.3) | 22.2 (16.1) |  |

Knowledge level: Low = 0-33.33%, Medium = 33.34-66.66%, High = 66.67-100%
$X^2(df)$ = Chi-square (degrees of freedom), $f$ = Fisher's exact p-value.

A comparison of the range of scores between ANC and delivery participants on knowledge about helminthiasis is presented in Figure 1. Though the median scores (<20%) at the two units (ANC–Delivery) appear similar and positively skewed, a bigger box, indicating a wider interquartile range was obtained at delivery. The range of values however were more varied at ANC than at delivery as more ANC participants had extremely high scores than those at delivery.

**Figure 1:**
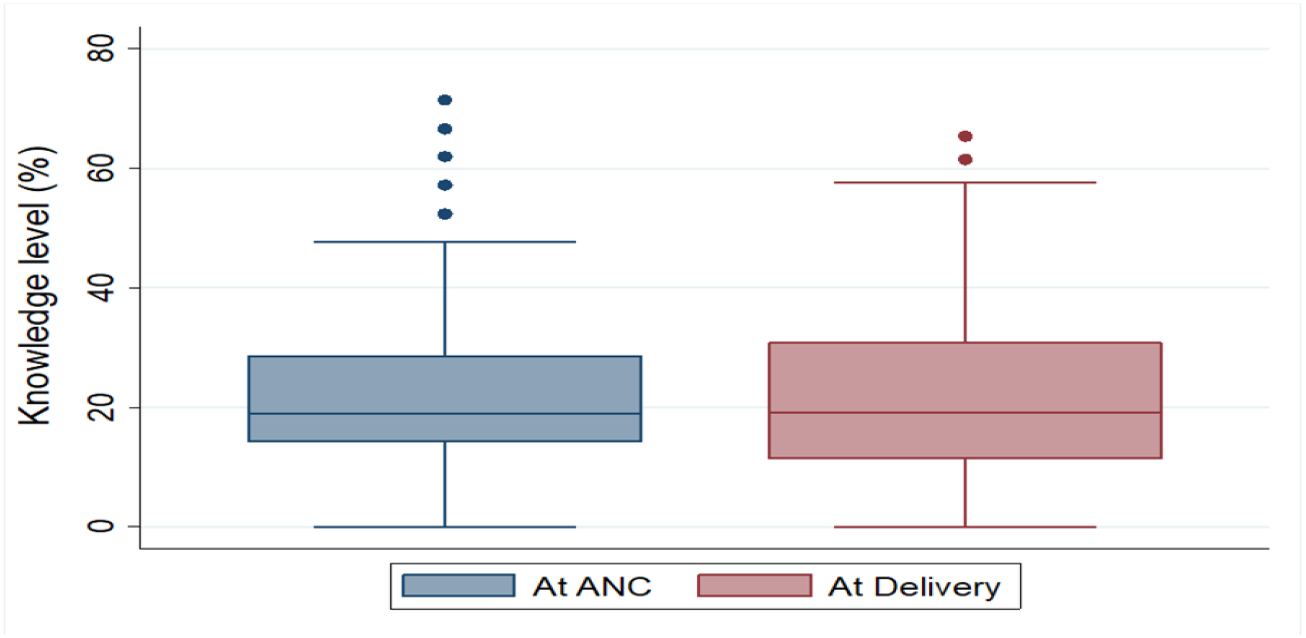
Comparison of general knowledge (% scale) at ANC and Delivery.

In Table 5 is the detailed summary of responses to questions on risks and prevention of helminth infections during pregnancy. The highest proportions of 69.2% in Battor and 27.3% in Adidome ticked anaemia as the risk of helminth infection during pregnancy. A much lower proportion (11.3% in Battor and 13.1% in Adidome) ticked ‘low birth weight’ (*X*^*2*^(16) =229.48, p<0.001). Participants who indicated ‘no knowledge’ were 38.6% in Adidome and 22.6% in Battor.

**Table 5:** Knowledge about the risk and prevention of helminths during pregnancy.

|  | ANC Health facility, N (%) |  | Test statistic | P-value |
| --- | --- | --- | --- | --- |
|  | Adidome | Battor |  |  |
| Knowledge about risks during pregnancy** | | | $X^2(16)=229.48$ | $p<0.001$ |
| Anaemia | 123 (27.3) | 263 (69.2) |  |  |
| Complications of the pregnancy | 95 (21.1) | 86 (22.6) |  |  |
| Low birth weight | 59 (13.1) | 43 (11.3) |  |  |
| Preterm birth | 39 (8.7) | 20 (5.3) |  |  |
| No knowledge | 174 (38.6) | 86 (22.6) |  |  |
| Knowledge about prevention during pregnancy** | | | $X^2(3)=4.34$ | $p=0.227$ |
| No knowledge | 341 (75.6) | 308 (81.1) |  |  |
| Taking Dewormer | 100 (22.2) | 68 (17.9) |  |  |
| Traditional/herbal prevention | 2 (0.4) | 1 (0.3) |  |  |
| Total | 451 (100.0) | 380 (100.0) |  |  |

\*\*Multiple responses allowed and so totals for variable may exceed totals below. $X^2(df)$ = Chi-square (degrees of freedom).

On the issue of knowledge about the prevention of helminths during pregnancy, a high proportion of participants (75.6% in Adidome and 81.1% in Battor) also indicated ‘no knowledge’. A small proportion of participants (0.4% in Adidome and 0.3% in Battor) however specified knowledge of ‘herbal preparations’ while (22.2% in Adidome and 17.9% in Battor) specified taking ‘dewormers’ as the prevention methods during pregnancy (*X*^*2*^(3) = 4.34, p = 0.227).

The questions on prevention of helminths in newborn babies got responses such as ‘Boiling/filtering drinking water’, ‘Exclusive breast feeding’, ‘Good personal hygiene’ and ‘Deworming every three months’ as presented in Table 6. In Adidome, only 5.3% and 3.0% stated ‘boiling/filtering drinking water’ and ‘Deworming every three months’ respectively as preventive measures for babies. Similarly, in Battor, a proportion of 12.9% and 3.1% participants respectively held the same view. Participants who stated, ‘Exclusive breast feeding’ were 14.6% in Adidome and 14.7% in Battor. ‘Good personal hygiene’ was chosen by 14.3% of participants in Adidome as against 9.2% participants in Battor (*X*^*2*^(31) = 50.94, p = 0.013).

**Table 6:** Knowledge about prevention of helminths in newborn babies.

|  | Delivery Health facility,<br>N (%) |  | Test statistic<br>P-value |
| --- | --- | --- | --- |
|  | Adidome | Battor |  |
| Knowledge about prevention of worms in babies** | | | $X^2(31) = 50.94$<br>$p = 0.013$ |
| Boiling/filtering drinking water | 16 (5.3) | 21 (12.9) |  |
| Exclusive breast feeding | 44 (14.6) | 24 (14.7) |  |
| Clean environment | 27 (9.0) | 9 (5.5) |  |
| Good personal hygiene | 43 (14.3) | 15 (9.2) |  |
| Eating properly cooked meals | 14 (4.7) | 2 (1.2) |  |
| Deworming every 3 months | 9 (3.0) | 5 (3.1) |  |
| Total | 301(100.0) | 163(100.0) |  |

\*\*Multiple responses allowed and so totals for variable may exceed totals below.
$X^2(df)$ = Chi-square (degrees of freedom).

Details of responses on general knowledge about the causes, symptoms and prevention of helminth infections are presented in Table 7. A general observation shows the highest proportion of participants both at ANC and delivery selected ‘vomiting’ as a symptom of helminth infection. Among ANC participants, the proportions were 33.9% in Adidome and 45.3% in Battor and among delivery participants, the proportions were 45.2% in Adidome and 45.4% in Battor. ‘Stomach ache’ and ‘diarrhoea’ were other common responses observed among participants as symptoms of helminth infections. Among ANC participants in Adidome, the proportions observed were 22.2% and 22.6% respectively. In Battor too, the proportions were 13,7% and 32.1% respectively. Among delivery participants in Adidome, the proportions observed for ‘Stomach ache’ and ‘diarrhoea’ were 38.5% and 22.3%, respectively and in Battor 53.4% and 23.3%, respectively, were observed.

**Table 7:** General knowledge on causes, symptoms and prevention of helminthiasis.

|  | ANC Health facility,<br>N (%) |  | Test statistic<br>P-value | Delivery Health facility,<br>N (%) |  | Test statistic<br>P-value |
| --- | --- | --- | --- | --- | --- | --- |
|  | Adidome | Battor |  | Adidome | Battor |  |
| Knowledge on symptoms of worm infections** | 102 (22.6) | 122 (32.1) | $X^2(37) = 111.57$<br>$p < 0.001$ | 67 (22.3) | 38 (23.3) | $X^2(47) = 107.49$<br>$p < 0.001$ |
| Diarrhoea | 70 (15.5) | 42 (11.1) |  | 14 (4.7) | 38 (23.3) |  |
| Headache | 153 (33.9) | 172 (45.3) |  | 136 (45.2) | 74 (45.4) |  |
| Vomiting | 100 (22.2) | 52 (13.7) |  | 116 (38.5) | 87 (53.4) |  |
| Stomach ache | 38 (8.4) | 32 (8.4) |  | 11 (3.7) | 31 (19.0) |  |
| Weakness | 67 (14.9) | 33 (8.7) |  | 82 (27.2) | 66 (40.5) |  |
| Loss of appetite |  |  |  |  |  |  |
| Knowledge on causes of worm infections** | 118 (26.2) | 229 (60.3) | $X^2(20) = 196.51$<br>$p < 0.001$ | 118 (39.2) | 80 (49.1) | $X^2(28) = 79.21$<br>$p < 0.001$ |
| Eating improperly cooked food | 80 (17.7) | 146 (38.4) |  | 119 (39.5) | 48 (29.6) |  |
| Playing in the sand | 86 (19.1) | 171 (45.0) |  | 104 (34.6) | 73 (44.8) |  |
| Poor sanitation | 67 (14.9) | 79 (20.8) |  | 103 (34.2) | 52 (31.9) |  |
| Exposure to human excreta | 5 (1.1) | 3 (0.8) |  | 9 (3.0) | 26 (16.0) |  |
| Malnutrition |  |  |  |  |  |  |
| Knowledge on prevention of worm infections** | 142 (31.5) | 218 (57.4) | $X^2(23) = 223.01$<br>$p < 0.001$ | 127 (42.2) | 80 (49.1) | $X^2(26) = 45.09$<br>$p = 0.011$ |
| Clean environment | 213 (47.2) | 150 (39.5) |  | 144 (47.8) | 69 (42.3) |  |
| Good personal hygiene | 8 (1.8) | 11 (2.9) |  | 10 (3.3) | 7 (4.3) |  |
| Sleeping under a bed net | 4 (0.9) | 3 (0.8) |  | 0 (0.0) | 0 (0.0) |  |
| Can't be prevented | 11 (2.4) | 36 (9.5) |  | 115 (38.2) | 59 (36.2) |  |
| Deworming every 3 months | 13 (2.9) | 110 (29.0) |  | 91 (30.2) | 66 (40.5) |  |
| Eating properly cooked meals |  |  |  |  |  |  |
| Total | 451 (100.0) | 380 (100.0) |  | 301 (100.0) | 163 (100.0) |  |

\*\*Multiple responses allowed and so totals for variable may exceed totals below. $X^2(df)$ = Chi-square (degrees of freedom).

General knowledge about the cause of helminth infections showed majority of Adidome participants at ANC (26.2%) and delivery (39.2%) ticked ‘eating improperly cooked food’. The highest proportion of participants who ticked ‘playing in the sand’ was observed among delivery participants in Adidome (39.5%) and the highest proportion who ticked ‘poor sanitation’ was observed among ANC participants in Battor.

General knowledge on prevention of helminth infections saw majority of the participants across both facilities and units ticking ‘clean environment’ and ‘good personal hygiene’. A smaller proportion of ANC participants (2.4% in Adidome and 9.5% in Battor) however chose “deworming every three months”. The corresponding proportion of delivery participants were 38.2% in Adidome and 36.2% in Battor.

Information in Table 8 is the mean scores on knowledge about helminthiasis across sociodemographic characteristics estimated as percentages of correct answers from the series of questions to determine participant knowledge. Across age categories, participants in Battor received higher scores than their counterparts in Adidome with the 40+ age category showing the highest mean score of 31.9 ± 11.5.

**Table 8:** Knowledge on helminths across sociodemographic characteristics.

|  | Knowledge level, Mean (SD) |  |
| --- | --- | --- |
|  | Adidome | Battor |
| <u>Age (years)</u> |  |  |
| <20 | 16.1 (11.3) | 19.9 (14.3) |
| 20 – 29 | 17.6 (10.5) | 24.9 (15.5) |
| 30 – 39 | 19.1 (10.7) | 29.1 (16.4) |
| 40+ | 16.7 (12.7) | 31.9 (11.5) |
| <u>Residence</u> |  |  |
| Adidome | 17.0 (10.8) | 20.2 (18.0) |
| Battor | 25.2 (8.4) | 27.3 (15.9) |
| Other | 17.4 (10.6) | 24.6 (15.5) |
| <u>Marital status</u> |  |  |
| Married | 18.1 (11.1) | 27.9 (15.4) |
| Single | 16.9 (10.3) | 19.2 (14.2) |
| Cohabiting | 19.0 (0.0) | 10.8 (12.2) |
| <u>Formal education</u> |  |  |
| None | 15.4 (10.1) | 21.8 (15.0) |
| Primary | 15.7 (10.4) | 21.0 (14.6) |
| Junior High School | 17.9 (10.6) | 24.3 (14.4) |
| Senior High School | 22.5 (10.6) | 29.0 (14.9) |
| Tertiary | 25.9 (16.0) | 42.9 (14.0) |
| <u>Gravida</u> |  |  |
| Primigravidae | 16.8 (10.6) | 24.6 (15.0) |
| Multigravidae | 18.1 (10.9) | 26.5 (16.0) |

Overall knowledge level scale: 0 (Lowest) to 100 (Highest).

A similar trend was observed across marital status where participants in Battor received higher scores than those in Adidome. Mean scores across all the other sociodemographic characteristics show participants in Battor with higher scores.

### Limitation

Correlation or association between knowledge of helminth infection among the study population and prevalences observed could not be performed due to the very low prevalences observed. There were only 10 PCR schistosomiasis positive cases, nine from Battor with only two having background information and one *Necator americanus* positive case so could not be used for comparative analysis with knowledge. The Kato Katz data also showed three schistosomiasis positive cases and one positive *Trichuris*. Thus, comparative analysis could not be performed as well.

## Discussion

The study set out to determine prevalence of intestinal helminths among pregnant women and compare local knowledge about helminthiasis in two rural communities where school children had been targets for helminth control to the total exclusion of the elderly and pregnant women.

### Prevalence of STH and *S. mansoni*

The low prevalence observed among study participants compares to prevalences observed among school children in a previous study (13) in some three districts in the Volta region. Even though, known risk factors for helminth infections such as exposure to contaminated fresh-water bodies, soil contact activities through farming and other agricultural practices were present in the study areas. The very low prevalence observed may be due to preventive measures such as public education on ‘clean environment’ and ‘personal hygiene’ to which a considerable proportion of the participants attested (Table 7).

### Deworming history and knowledge about dewormers

Although a high percentage of study participants reportedly do not regularly take dewormers (Table 2), that did not reflect in the prevalence of helminthic infection observed. Other factors not identified by the study might account for the low prevalence. Since deworming helps to reduce morbidity in infected individuals, there is the need for public education on the importance of periodic deworming most especially for the asymptomatic carriers.

### Local knowledge about causes, symptoms and prevention of helminth infections

The overall low level of knowledge observed (Table 4 and Figure 1) coupled with other existing predisposing factors indicated that the study populations were at risk of helminth infections. This observation was confirmed by the observation that a high percentage of participants indicated no knowledge of preventive measures during pregnancy (Table 5). This low-level knowledge could also account for the study populations’ dislike for deworming as a preventive measure. Battor as a community has more schools than Adidome. Consequently, it was expected of participants from Battor to show better knowledge about helminthiasis as has been observed in Table 4.

Common perception of risk of helminth infection among study participants were anemia, pregnancy complications and low birth weight.

## Conclusion

In spite of the low prevalence observed, the study revealed low knowledge about helminthiasis among the study population. The cause for the low prevalence of helminth infection among study participants could not be identified. Intensifying health education as community-based interventions is still necessary for the effective control and elimination of schistosomiasis and soil transmitted helminths in the study area. There is also the need to include pregnant women in future control programs to achieve complete elimination of intestinal helminths.

## Data Availability

There are no restrictions to data sharing

## Acknowledgements

Sincere thanks and gratitude to IRD (Institut de Recherche pour le Développement), funders of this study and to Mr. Tony Godi (School of Public Health, University of Ghana) for the analysis of the data. Our deepest appreciation to the staff of Battor Catholic Hospital and the staff of Adidome Government Hospital for all their assistance.

